# Differential expression of genes influencing mitotic processes in cord blood mononuclear cells after a pre-conceptional micronutrient-based randomized controlled trial: Pune Rural Intervention in Young Adolescents (PRIYA)

**DOI:** 10.1101/2021.08.25.21262585

**Authors:** Satyajeet P. Khare, Ayush Madhok, Indumathi Patta, Krishna K. Sukla, Vipul V. Wagh, Pooja S. Kunte, Deepa Raut, Dattatray Bhat, Kalyanaraman Kumaran, Caroline Fall, Utpal Tatu, Giriraj R. Chandak, Chittaranjan S. Yajnik, Sanjeev Galande

**Author notes:** These authors contributed equally (Satyajeet P. Khare, Ayush Madhok).

## Abstract

In The Pune Maternal Nutrition Study, vitamin B12 deficiency was seen in 65% of pregnant women, folate deficiency was rare. Maternal total homocysteine concentrations were inversely associated with offspring birthweight, and low vitamin B12 and high folate concentrations predicted higher offspring adiposity and insulin resistance. These findings guided a nested pre-conceptional randomized controlled trial ‘Pune Rural Intervention in Young Adolescents (PRIYA)’. The interventions included: 1) vitamin B12+multi-micronutrients the United Nations International Multiple Micronutrient Antenatal Preparation (UNIMMAP) and proteins (B12+MMN), 2) vitamin B12 (B12 alone), and 3) placebo. Intervention improved maternal pre-conceptional and in-pregnancy micronutrient nutrition. Gene expression analysis in cord blood mononuclear cells in 88 pregnancies revealed 75 differentially expressed genes between the B12+MMN and placebo groups. The enriched biological processes included G2/M phase transition, chromosome segregation, and nuclear division. Enriched pathways included, mitotic spindle checkpoint and DNA damage response while enriched human phenotypes were sloping forehead and decreased head circumference. Fructose-bisphosphatase 2 (*FBP2*) and Cell Division Cycle Associated 2 (*CDCA2*) genes were under-expressed in the B12 alone group. The latter, involved in chromosome segregation was under-expressed in both intervention groups. Based on the role of B-complex vitamins in the synthesis of nucleotides and S-Adenosyl Methionine, and the roles of vitamins A and D on gene expression, we propose that the multi-micronutrient intervention epigenetically affected cell cycle dynamics. Neonates in the B12+MMN group had the highest ponderal index. Follow up studies will reveal if the intervention and the altered biological processes influence offspring diabesity.

## Introduction

The Developmental Origins of Health and Disease (DOHaD) paradigm proposes that the environment during crucial stages of development modifies gene expression through epigenetic influences which permanently affect the structure and function of the developing organism^1^. This phenomenon is also called ‘fetal programming’^2^. This paradigm originated from Prof. David Barker’s demonstration that low birth weight (indicative of poor fetal nutrition) was a risk factor for future diabetes and cardiovascular disease^3^. DOHaD theory proposes that improving fetal nutrition will help prevent common non-communicable diseases.

Fetal programming is largely an epigenetic phenomenon and depends on chemical modifications of DNA and histones or action of microRNAs, all of which influence gene expression^4^. DNA methylation is the most studied mechanism and is influenced by dietary nutrients which provide or generate methyl groups for cellular metabolism^5, 6^. These nutrients include vitamins B12, B2, B6, folate, choline, betaine, and proteins^7^. Vitamins A and D are also known to influence gene expression through their interaction with the nuclear receptors^8, 9^. Deficiency of these nutrients in the diet of a pregnant mother manifests in adverse effects on the fetus, including developmental defects (for example, neural tube defects), growth abnormalities (growth restriction)^10^, and are also associated with an increased risk of non-communicable diseases in later life^11, 12^.

The Pune Maternal Nutrition Study (PMNS) is a community based pre-conceptional birth cohort started in 1993 based on Prof. Barker’s theory^13^. It was set up in six villages near Pune to study maternal nutritional determinants of fetal growth and to study life-course evolution of the risk of diabetes and related traits in the offspring. We found that two thirds of the pregnant mothers had low vitamin B12 status but folate status was adequate in the majority^14^. Lower maternal folate and higher total homocysteine concentrations were associated with lower offspring birth weight^13, 15^. Low maternal vitamin B12 and normal to high folate concentrations were associated with increased risk of diabesity (insulin resistance and adiposity), and low maternal vitamin D status with higher adiposity in the offspring^14, 16^. These findings prompted us to set up the Pune Rural Intervention in Young Adolescents (PRIYA) trial^17^. The trial provided vitamin B12 alone or along with other micronutrients (B-complex vitamins, vitamins A, D, and E, and trace elements) and milk proteins to the adolescent women in the PMNS cohort. The long-term aim of this intervention is to reduce diabesity and improve neurocognition in their offspring. In the short term, we aimed to investigate changes in DNA methylome, transcriptome, proteome, and metabolome in the cord blood of the newborn and its birth size. We have recently reported better neurocognitive outcomes in young offspring in the PRIYA trial whose mothers received vitamin B12 alone^18^.

The purpose of this paper is to describe the transcriptomic changes in the cord blood which may provide a mechanistic insight into micronutrient-mediated fetal programming, and possible pathways which may influence long-term outcomes in the offspring.

## Materials and Methods

### The PRIYA trial - Ethics permissions and trial registration

PRIYA is a randomized, double blind, placebo-controlled trial in adolescent participants of the PMNS (Figure 1). The rationale and methods of the PRIYA trial are described previously^17^. The PRIYA trial was approved by the KEM Hospital Research Centre Ethics committee and monitored by a Data Safety Monitoring Board (DSMB) and a Scientific Advisory Committee (SAC). Informed consent was signed by parents of all participants, and an informed assent was signed by participants below 18 years of age. After attaining 18 years of age, all participants signed their own informed consent. The trial was registered with the CTRI (2012/12/003212) and ISRCTN (32921044).

**Figure 1:**
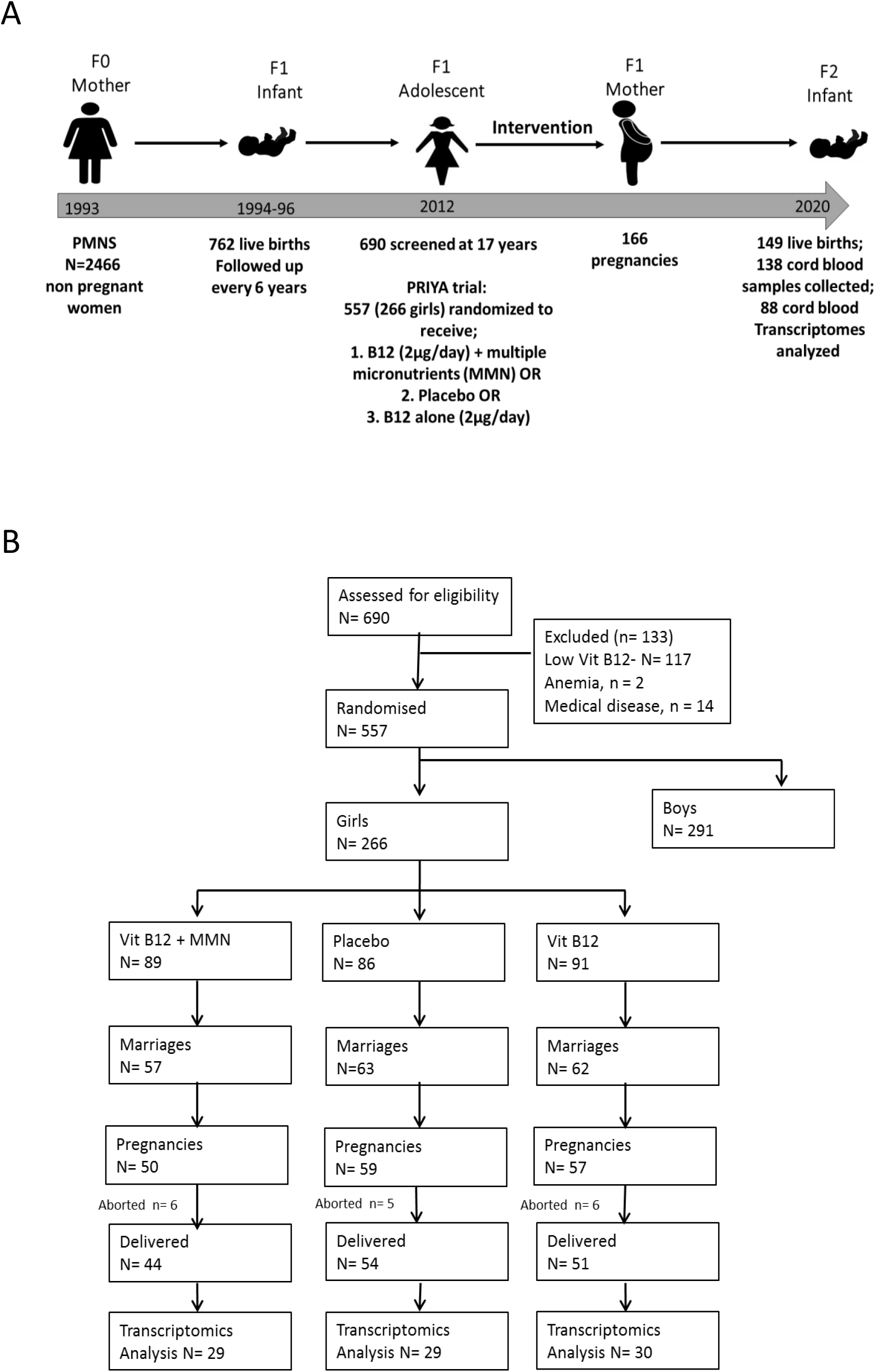
The Pune Maternal Nutrition Study (PMNS) and the Pune Rural Intervention in Young Adolescents (PRIYA) trial. **A)** Broad plan of the study. B) CONSORT diagram of the PRIYA trial.

### Participants

Adolescent participants in the PMNS study were screened for vitamin B12 levels and hemoglobin concentrations. Those with severe vitamin B12 deficiency (plasma vitamin B12 concentration <100 pmol/L) or severe anaemia (blood hemoglobin concentration <70 gm/L) were excluded from the trial and treated appropriately. This was to fulfil ethical imperatives of a placebo-controlled trial. Those with chronic medical conditions were also excluded.

### Intervention and Comparator

Eligible participants were randomised into 3 intervention groups: 1) vitamin B12 (2 μg/day) along with multi-micronutrients as per United Nations International Multiple Micronutrient Antenatal Preparation (UNIMMAP)^19^, excluding folic acid and conforming to local regulatory requirements (Supplementary Table 1), 2) Placebo, 3) vitamin B12 (2 μg/day) alone. The multi-micronutrient group also received 20 gm of milk powder every day and all 3 groups received iron and folic acid as per Government of India approved dosage (60 mg elemental iron + 500 μg folic acid once a week before pregnancy and one tablet daily for at least 100 days after diagnosis of pregnancy). Every month, bottles were distributed by local field staff who were in regular touch with the participants. Bottles were returned each month for counting the unused capsules, this information was used for calculating compliance. The trial intervention was continued until delivery of the first live child.

Habitual dietary intake was assessed using food frequency questionnaire (FFQ), developed and validated in the PMNS^13^.

### Outcomes

A fasting blood sample was obtained for measurement of circulating micronutrients 6-12 months after the intervention was started. Marriages were recorded, and pregnancies were confirmed by a urine pregnancy test. At 28-weeks’ gestation women visited KEM Hospital for an obstetric consultation and blood tests (hemogram, micronutrient levels and a fasting 75g oral glucose tolerance test). In the villages, women consulted local medical practitioners as required. The field staff visited and monitored pregnant women regularly and helped transfer them for delivery at the King Edward Memorial (KEM) Hospital, Pune.

Trained research staff attended all deliveries and collected maternal and cord blood samples as per a standardised protocol. An aliquot of cord blood was transferred to PAXgene tubes (BD, USA) and stored at -80°C till processed for extraction of RNA. Details of the delivery were recorded. Detailed anthropometric measurements were made within 24 hours of delivery by trained research staff using standardised methods (Supplementary Table 2). Gestational age was calculated from last menstrual period and confirmed by ultrasound measurements, if required.

### Blood measurements

Maternal plasma vitamin B12, B6, holo-TC (‘active’ vitamin B12), folate, and total homocysteine concentrations were measured at screening, 6-12 months after starting intervention, at 28 weeks gestation, and at delivery. Similar measurements were made in the cord blood. Vitamin B2 was measured at all time-points except the screening. Supplementary Table 2 describes the laboratory methods used for these measurements.

### Statistical methods for clinical characteristics

Maternal and neonatal clinical characteristics are shown as median (5^th^-95^th^ centiles). The significance of difference between intervention and placebo groups was tested by Mann-Whitney test.

### Isolation of cord blood mononuclear cells

Mononuclear cells from the cord blood were extracted after depleting RBCs from whole blood. In brief, batches of frozen blood samples were thawed and diluted with one volume of phosphate buffer saline supplemented with EDTA (2mM). One volume of RBC lysis buffer (Biolegend # 420301) was added to the diluted samples and incubated for 5 minutes at RT. The tubes were centrifuged at *350 x g* for 15 min at 4°C and the pellets were processed for RNA isolation.

### RNA isolation

Cord blood mononuclear cells were subjected to RNA isolation using TriZol reagent (Invitrogen). Quantitation of RNA was performed on Qubit 4 Fluorometer (Thermo Scientific #Q33238) using RNA HS Kit (Thermo Scientific #Q32852). Libraries were prepared according to the manufacturer’s protocol (Illumina). Briefly, each sample was run on 2100 Bioanalyzer (Agilent #G2939BA) to check its integrity. About 500 ng of total RNA from each sample was used to make libraries using TruSeq Stranded mRNA Sample Prep Kit (Illumina #20020594). After final quantification and pooling, the libraries were run on Illumina HiSeqX platform, with 150 nucleotides long paired end reads as the output.

### RNA sequencing and data analysis

The quality of sequencing was assessed using FastQC (v0.11.8). The raw sequence reads were aligned to the human genome (hg38, GENCODE)^20^ using HiSAT2^21^. The resulting BAM files were used to generate a count matrix using featureCounts for known genes^22^. Samples with reads that aligned poorly (Q1-IQR*1.5) were excluded from the downstream analysis. Low expression genes (less than 20 counts in total in all samples) were not included for the differential expression analysis. Gene expression levels were normalized by internal normalization method by calculating geometric mean in DESEq2^23^. Normalized expression values were regularized log transformed for PCA plots. Differential expression analysis was performed between the placebo group and the intervention groups by two-group comparison using the default Wald test in DESeq2. Maternal age at the start of intervention, duration of intervention, sex of the child, and cord blood monocyte, granulocyte, and lymphocyte counts were used as covariates. Absolute values of these covariates were converted to categorical variables around the median. Since the samples were processed in batches, the batch numbers were also used as a covariate. FDR adjusted P-value cut-off of 0.05 and base mean value cut-offs of 1 were used for identification of differentially expressed genes. Significantly differentially expressed genes were subjected to gene enrichment analysis using clusterProfiler (v3.14.3)^24^ and gProfiler (version:e105_eg52_p16_e84549f)^25^. To understand the gene-networking, the differentially expressed genes (DEGs), were subjected to gene network analysis using STRING^26^.

## Results

### Trial progress and clinical findings

Six hundred and ninety adolescent participants in the PMNS (362 boys, 328 girls) were screened for inclusion in the PRIYA trial in the year 2012 (Figure 1). A total of 133 were excluded and 557 (266 females, 291 males) were randomized into 3 groups. The intervention was started in September 2012.

This paper refers to outcomes only in female participants. At screening they were ∼16 years old, had a median body mass index (BMI) of 18.0 kg/m^2^ (56.7% had a BMI <18.5 kg/m^2^, 30.1% were anemic (Hemoglobin <120 gm/L), and average education was 13 years of schooling. This is a predominantly vegetarian population: a third never ate non-vegetarian foods, and a fourth ate it less than once a week. Portion sizes of non-vegetarian foods were very small. Only 30% consumed milk more than alternate days. A total of 182 females married during the trial, 166 became pregnant, and 149 delivered in the study. Cord blood samples were available on 138 for gene expression analysis. This paper reports transcriptome measurements on the first 88 cord blood samples (40 girls, 48 boys); these pregnancies were comparable to the 50 who are not included in this study [for maternal age, BMI, socio-economic status, and hemoglobin concentrations at 28 weeks gestation, p > 0.05].

At 28 weeks gestation, the mothers were on average 21 years old and weighed 53.0 kgs with a BMI of 22.0 kg/m^2^ (Table 1). The first delivery was in November 2014 and the last delivery was in February 2020. The duration of intervention was on an average of 54 months (min = 24, max = 83). Three mothers delivered pre-term, and 23 were delivered by caesarean section. The average birth weight of babies was 2.8 kg, 14 were low birth weight (<2.5 kg) (Table 2). There was no significant difference in the birth weight, length, skin fold thicknesses in the 3 groups; however, ponderal index was highest in the B12+MMN group. The pregnancy outcomes (pre-term deliveries, caesarean sections, and common adverse events) were similar in the three intervention groups.

**Table 1:**
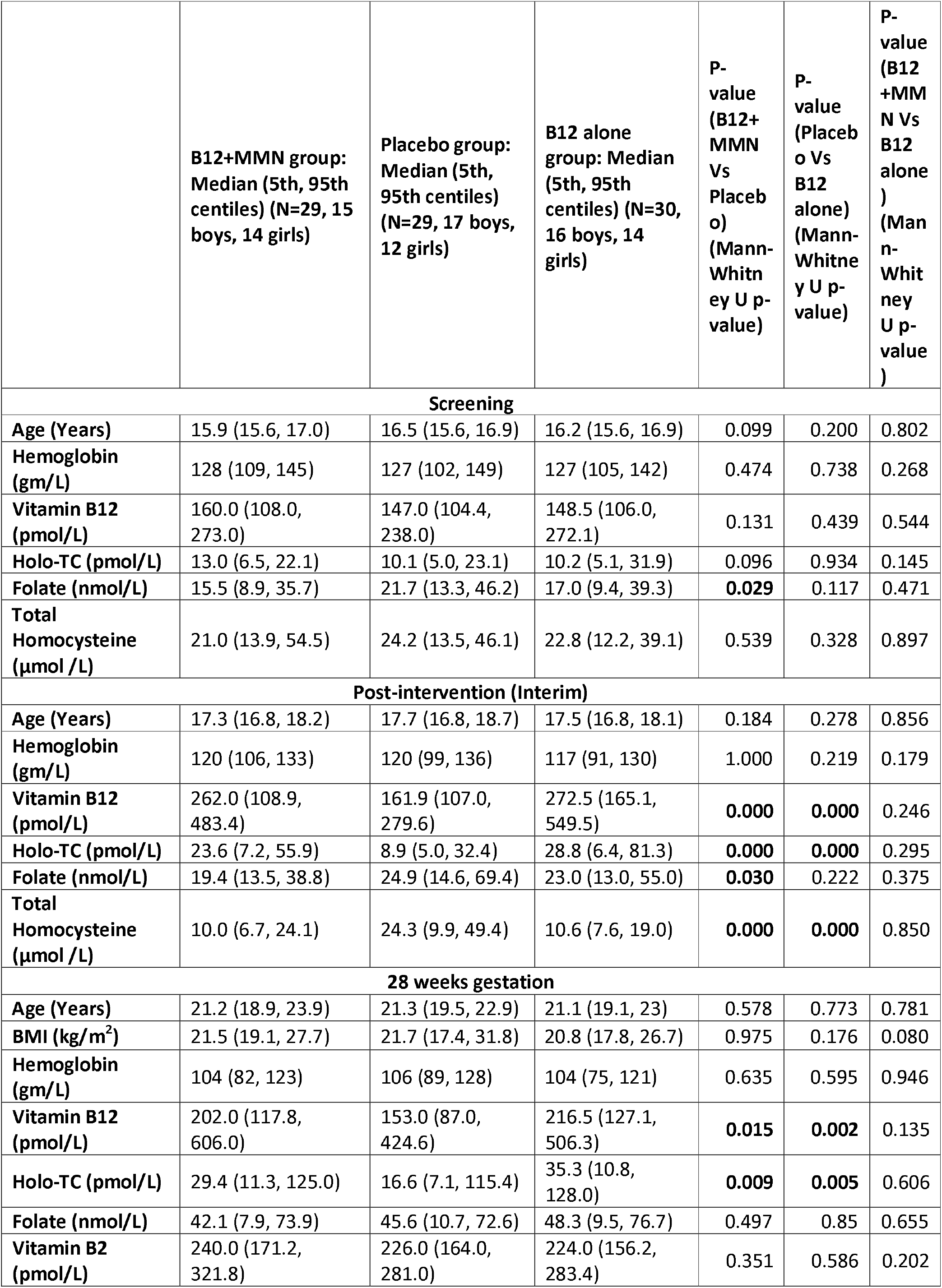
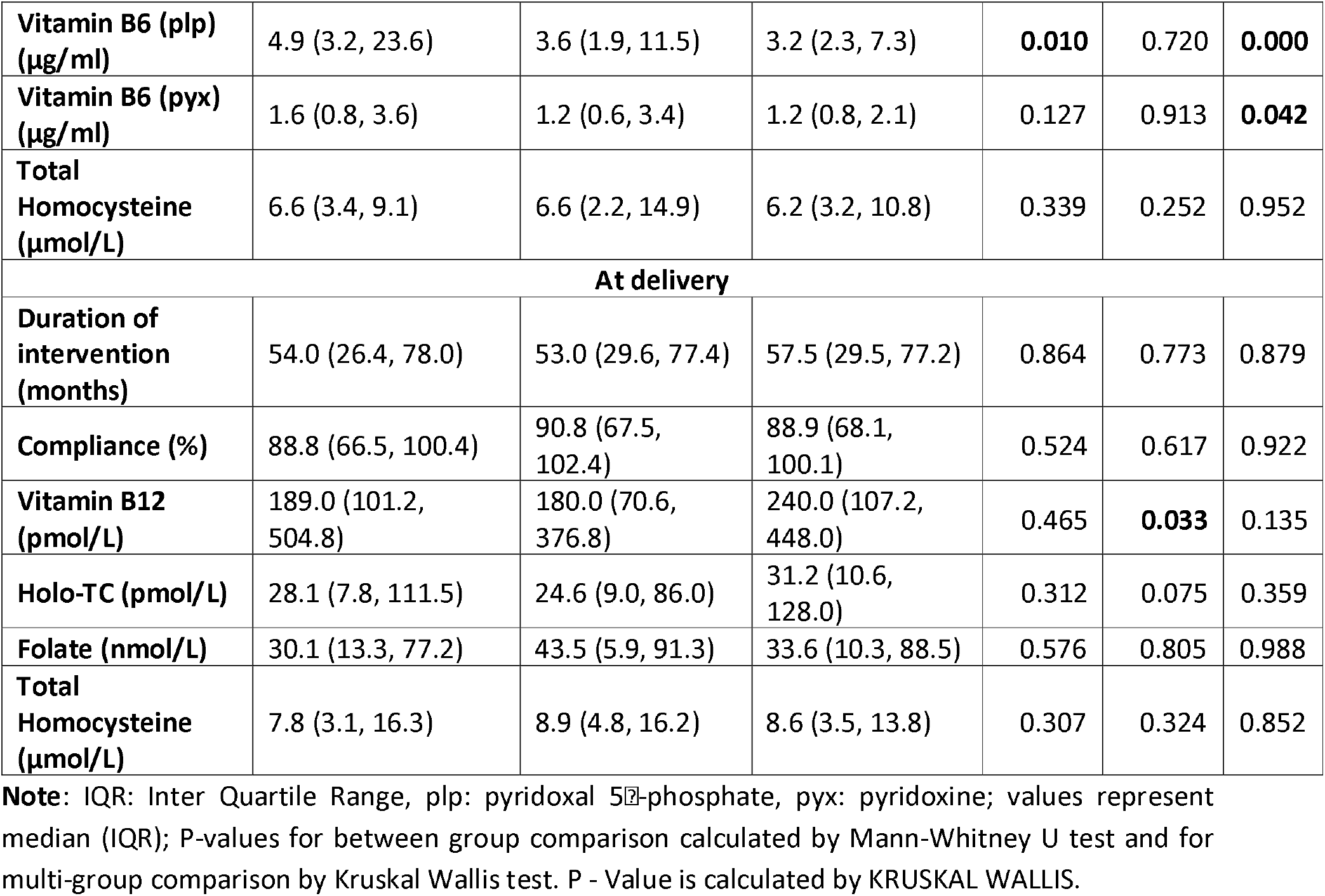
Comparison of maternal characteristics between study participants of 3 intervention groups.

|  | **B12+MMN group:**<br>**Median (5th, 95th**<br>**centiles) (N=29, 15**<br>**boys, 14 girls)** | **Placebo group:**<br>**Median (5th, 95th**<br>**centiles) (N=29, 17 boys,**<br>**12 girls)** | **B12 alone**<br>**group: Median**<br>**(5th, 95th**<br>**centiles) (N=30,**<br>**16 boys, 14**<br>**girls)** | **P-**<br>**value**<br>**(B12+**<br>**MMN**<br>**Vs**<br>**Placebo)**<br>**(Mann-**<br>**Whitney U**<br>**p-value)** | **P-**<br>**value**<br>**(Placebo**<br>**Vs**<br>**B12**<br>**alone)**<br>**(Mann-**<br>**Whitney U**<br>**p-value)** | **P-**<br>**value**<br>**(B12**<br>**+MMN**<br>**Vs**<br>**B12**<br>**alone)**<br>**(Mann-**<br>**Whitney U**<br>**p-value)** |
| --- | --- | --- | --- | --- | --- | --- |
| **Screening** |  |  |  |  |  |  |
| **Age (Years)** | 15.9 (15.6, 17.0) | 16.5 (15.6, 16.9) | 16.2 (15.6, 16.9) | 0.099 | 0.200 | 0.802 |
| **Hemoglobin**<br>**(gm/L)** | 128 (109, 145) | 127 (102, 149) | 127 (105, 142) | 0.474 | 0.738 | 0.268 |
| **Vitamin B12**<br>**(pmol/L)** | 160.0 (108.0,<br>273.0) | 147.0 (104.4,<br>238.0) | 148.5 (106.0,<br>272.1) | 0.131 | 0.439 | 0.544 |
| **Holo-TC (pmol/L)** | 13.0 (6.5, 22.1) | 10.1 (5.0, 23.1) | 10.2 (5.1, 31.9) | 0.096 | 0.934 | 0.145 |
| **Folate (nmol/L)** | 15.5 (8.9, 35.7) | 21.7 (13.3, 46.2) | 17.0 (9.4, 39.3) | **0.029** | 0.117 | 0.471 |
| **Total**<br>**Homocysteine**<br>**(μmol /L)** | 21.0 (13.9, 54.5) | 24.2 (13.5, 46.1) | 22.8 (12.2, 39.1) | 0.539 | 0.328 | 0.897 |
| **Post-intervention (Interim)** |  |  |  |  |  |  |
| **Age (Years)** | 17.3 (16.8, 18.2) | 17.7 (16.8, 18.7) | 17.5 (16.8, 18.1) | 0.184 | 0.278 | 0.856 |
| **Hemoglobin**<br>**(gm/L)** | 120 (106, 133) | 120 (99, 136) | 117 (91, 130) | 1.000 | 0.219 | 0.179 |
| **Vitamin B12**<br>**(pmol/L)** | 262.0 (108.9,<br>483.4) | 161.9 (107.0,<br>279.6) | 272.5 (165.1,<br>549.5) | **0.000** | **0.000** | 0.246 |
| **Holo-TC (pmol/L)** | 23.6 (7.2, 55.9) | 8.9 (5.0, 32.4) | 28.8 (6.4, 81.3) | **0.000** | **0.000** | 0.295 |
| **Folate (nmol/L)** | 19.4 (13.5, 38.8) | 24.9 (14.6, 69.4) | 23.0 (13.0, 55.0) | **0.030** | 0.222 | 0.375 |
| **Total**<br>**Homocysteine**<br>**(μmol /L)** | 10.0 (6.7, 24.1) | 24.3 (9.9, 49.4) | 10.6 (7.6, 19.0) | **0.000** | **0.000** | 0.850 |
| **28 weeks gestation** |  |  |  |  |  |  |
| **Age (Years)** | 21.2 (18.9, 23.9) | 21.3 (19.5, 22.9) | 21.1 (19.1, 23) | 0.578 | 0.773 | 0.781 |
| **BMI (kg/m<sup>2</sup>)** | 21.5 (19.1, 27.7) | 21.7 (17.4, 31.8) | 20.8 (17.8, 26.7) | 0.975 | 0.176 | 0.080 |
| **Hemoglobin**<br>**(gm/L)** | 104 (82, 123) | 106 (89, 128) | 104 (75, 121) | 0.635 | 0.595 | 0.946 |
| **Vitamin B12**<br>**(pmol/L)** | 202.0 (117.8,<br>606.0) | 153.0 (87.0,<br>424.6) | 216.5 (127.1,<br>506.3) | **0.015** | **0.002** | 0.135 |
| **Holo-TC (pmol/L)** | 29.4 (11.3, 125.0) | 16.6 (7.1, 115.4) | 35.3 (10.8,<br>128.0) | **0.009** | **0.005** | 0.606 |
| **Folate (nmol/L)** | 42.1 (7.9, 73.9) | 45.6 (10.7, 72.6) | 48.3 (9.5, 76.7) | 0.497 | 0.85 | 0.655 |
| **Vitamin B2**<br>**(pmol/L)** | 240.0 (171.2,<br>321.8) | 226.0 (164.0,<br>281.0) | 224.0 (156.2,<br>283.4) | 0.351 | 0.586 | 0.202 |
| Intergenerational effect of maternal intervention |  |  |  |  |  |  |
| **Vitamin B6 (plp) (µg/ml)** | 4.9 (3.2, 23.6) | 3.6 (1.9, 11.5) | 3.2 (2.3, 7.3) | **0.010** | 0.720 | **0.000** |
| **Vitamin B6 (pyx) (µg/ml)** | 1.6 (0.8, 3.6) | 1.2 (0.6, 3.4) | 1.2 (0.8, 2.1) | 0.127 | 0.913 | **0.042** |
| **Total Homocysteine (µmol/L)** | 6.6 (3.4, 9.1) | 6.6 (2.2, 14.9) | 6.2 (3.2, 10.8) | 0.339 | 0.252 | 0.952 |
| **At delivery** |  |  |  |  |  |  |
| **Duration of intervention (months)** | 54.0 (26.4, 78.0) | 53.0 (29.6, 77.4) | 57.5 (29.5, 77.2) | 0.864 | 0.773 | 0.879 |
| **Compliance (%)** | 88.8 (66.5, 100.4) | 90.8 (67.5, 102.4) | 88.9 (68.1, 100.1) | 0.524 | 0.617 | 0.922 |
| **Vitamin B12 (pmol/L)** | 189.0 (101.2, 504.8) | 180.0 (70.6, 376.8) | 240.0 (107.2, 448.0) | 0.465 | **0.033** | 0.135 |
| **Holo-TC (pmol/L)** | 28.1 (7.8, 111.5) | 24.6 (9.0, 86.0) | 31.2 (10.6, 128.0) | 0.312 | 0.075 | 0.359 |
| **Folate (nmol/L)** | 30.1 (13.3, 77.2) | 43.5 (5.9, 91.3) | 33.6 (10.3, 88.5) | 0.576 | 0.805 | 0.988 |
| **Total Homocysteine (µmol/L)** | 7.8 (3.1, 16.3) | 8.9 (4.8, 16.2) | 8.6 (3.5, 13.8) | 0.307 | 0.324 | 0.852 |

**Note:** IQR: Inter Quartile Range, plp: pyridoxal 5'-phosphate, pyx: pyridoxine; values represent median (IQR); P-values for between group comparison calculated by Mann-Whitney U test and for multi-group comparison by Kruskal Wallis test. P - Value is calculated by KRUSKAL WALLIS.

**Table 2:**
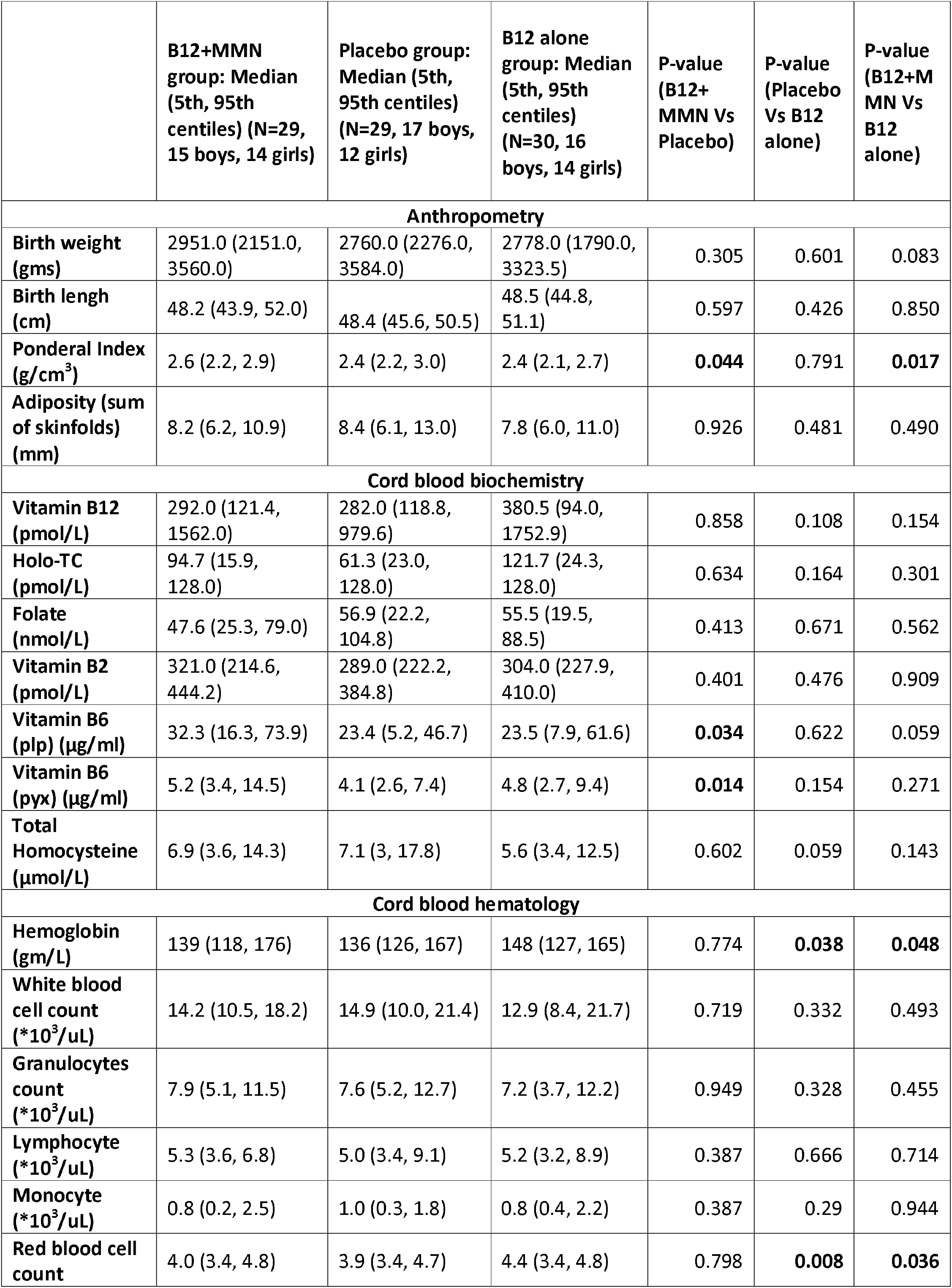
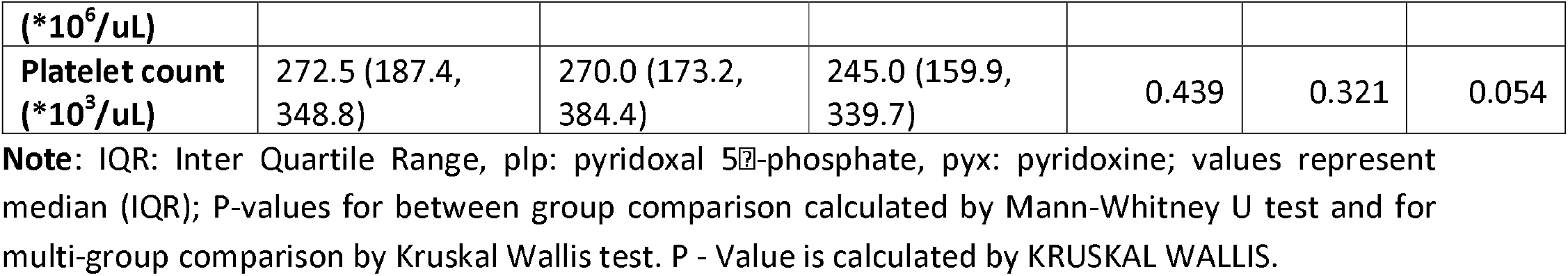
Comparison of neonatal characteristics between study participants of 3 intervention groups.

|  | **B12+MMN**<br>**group: Median**<br>**(5th, 95th**<br>**centiles) (N=29,**<br>**15 boys, 14 girls)** | **Placebo group:**<br>**Median (5th,**<br>**95th centiles)**<br>**(N=29, 17 boys,**<br>**12 girls)** | **B12 alone**<br>**group: Median**<br>**(5th, 95th**<br>**centiles)**<br>**(N=30, 16**<br>**boys, 14 girls)** | **P-value**<br>**(B12+**<br>**MMN Vs**<br>**Placebo)** | **P-value**<br>**(Placebo**<br>**Vs B12**<br>**alone)** | **P-value**<br>**(B12+M**<br>**MN Vs**<br>**B12**<br>**alone)** |
| --- | --- | --- | --- | --- | --- | --- |
| **Anthropometry** |  |  |  |  |  |  |
| **Birth weight**<br>**(gms)** | 2951.0 (2151.0,<br>3560.0) | 2760.0 (2276.0,<br>3584.0) | 2778.0 (1790.0,<br>3323.5) | 0.305 | 0.601 | 0.083 |
| **Birth length**<br>**(cm)** | 48.2 (43.9, 52.0) | 48.4 (45.6, 50.5) | 48.5 (44.8,<br>51.1) | 0.597 | 0.426 | 0.850 |
| **Ponderal Index**<br>**(g/cm<sup>3</sup>)** | 2.6 (2.2, 2.9) | 2.4 (2.2, 3.0) | 2.4 (2.1, 2.7) | **0.044** | 0.791 | **0.017** |
| **Adiposity (sum**<br>**of skinfolds)**<br>**(mm)** | 8.2 (6.2, 10.9) | 8.4 (6.1, 13.0) | 7.8 (6.0, 11.0) | 0.926 | 0.481 | 0.490 |
| **Cord blood biochemistry** |  |  |  |  |  |  |
| **Vitamin B12**<br>**(pmol/L)** | 292.0 (121.4,<br>1562.0) | 282.0 (118.8,<br>979.6) | 380.5 (94.0,<br>1752.9) | 0.858 | 0.108 | 0.154 |
| **Holo-TC**<br>**(pmol/L)** | 94.7 (15.9,<br>128.0) | 61.3 (23.0,<br>128.0) | 121.7 (24.3,<br>128.0) | 0.634 | 0.164 | 0.301 |
| **Folate**<br>**(nmol/L)** | 47.6 (25.3, 79.0) | 56.9 (22.2,<br>104.8) | 55.5 (19.5,<br>88.5) | 0.413 | 0.671 | 0.562 |
| **Vitamin B2**<br>**(pmol/L)** | 321.0 (214.6,<br>444.2) | 289.0 (222.2,<br>384.8) | 304.0 (227.9,<br>410.0) | 0.401 | 0.476 | 0.909 |
| **Vitamin B6**<br>**(plp) (µg/ml)** | 32.3 (16.3, 73.9) | 23.4 (5.2, 46.7) | 23.5 (7.9, 61.6) | **0.034** | 0.622 | 0.059 |
| **Vitamin B6**<br>**(pyx) (µg/ml)** | 5.2 (3.4, 14.5) | 4.1 (2.6, 7.4) | 4.8 (2.7, 9.4) | **0.014** | 0.154 | 0.271 |
| **Total**<br>**Homocysteine**<br>**(µmol/L)** | 6.9 (3.6, 14.3) | 7.1 (3, 17.8) | 5.6 (3.4, 12.5) | 0.602 | 0.059 | 0.143 |
| **Cord blood hematology** |  |  |  |  |  |  |
| **Hemoglobin**<br>**(gm/L)** | 139 (118, 176) | 136 (126, 167) | 148 (127, 165) | 0.774 | **0.038** | **0.048** |
| **White blood**<br>**cell count**<br>**(\*10<sup>3</sup>/uL)** | 14.2 (10.5, 18.2) | 14.9 (10.0, 21.4) | 12.9 (8.4, 21.7) | 0.719 | 0.332 | 0.493 |
| **Granulocytes**<br>**count**<br>**(\*10<sup>3</sup>/uL)** | 7.9 (5.1, 11.5) | 7.6 (5.2, 12.7) | 7.2 (3.7, 12.2) | 0.949 | 0.328 | 0.455 |
| **Lymphocyte**<br>**(\*10<sup>3</sup>/uL)** | 5.3 (3.6, 6.8) | 5.0 (3.4, 9.1) | 5.2 (3.2, 8.9) | 0.387 | 0.666 | 0.714 |
| **Monocyte**<br>**(\*10<sup>3</sup>/uL)** | 0.8 (0.2, 2.5) | 1.0 (0.3, 1.8) | 0.8 (0.4, 2.2) | 0.387 | 0.29 | 0.944 |
| **Red blood cell**<br>**count** | 4.0 (3.4, 4.8) | 3.9 (3.4, 4.7) | 4.4 (3.4, 4.8) | 0.798 | **0.008** | **0.036** |
| (*10<sup>6</sup>/uL) |  |  |  |  |  |  |
| **Platelet count**<br>(*10<sup>3</sup>/uL) | 272.5 (187.4,<br>348.8) | 270.0 (173.2,<br>384.4) | 245.0 (159.9,<br>339.7) | 0.439 | 0.321 | 0.054 |

**Note:** IQR: Inter Quartile Range, plp: pyridoxal 5'-phosphate, pyx: pyridoxine; values represent median (IQR); P-values for between group comparison calculated by Mann-Whitney U test and for multi-group comparison by Kruskal Wallis test. P - Value is calculated by KRUSKAL WALLIS.

At screening, plasma vitamin B12 concentrations were low and 47.7% were vitamin B12 deficient (<150 pmol/L), only 1% were folate deficient (<7 nmol/L). Plasma total homocysteine concentrations were high, 91% had hyperhomocysteinemia (>15 μmol/L). There was no significant difference in plasma vitamin B12, folate, and total homocysteine concentrations in the 3 intervention groups. In women, in the two vitamin B12 supplemented groups there was a sizeable rise in vitamin B12 and holo-TC concentrations compared to the placebo group, and a fall in total homocysteine concentrations within a few months (Figure 2A). The levels of vitamin B12 and total homocysteine fell during pregnancy (even in the placebo group), but both vitamin B12 and holo-TC remained higher in the two intervention groups. The difference in total homocysteine concentrations between groups was blunted in pregnancy. Vitamins B2 and B6 were the highest in the B12+MMN group at all time points after intervention (Table 1). Maternal hemoglobin concentrations were similar in the three groups throughout the trial. The measured compliance for consumption of intervention capsules was similar in the three intervention groups.

**Figure 2:**
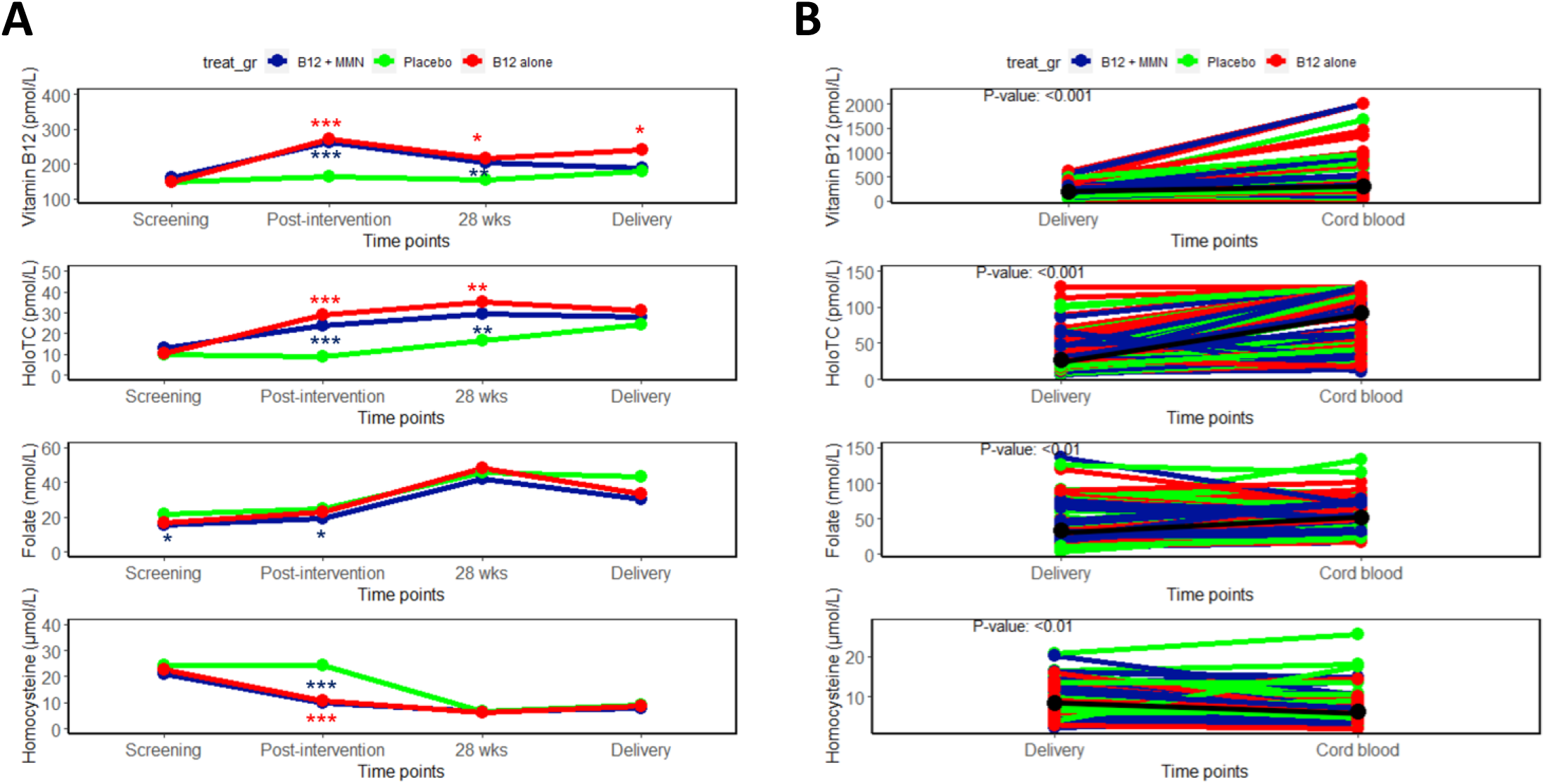
Serial micronutrient levels in the mothers and baby’s cord blood in the PRIYA trial. A) Median concentrations of vitamin B12 (pmol/L), Holo-TC (pmol/L), folate (nmol/L), and total homocysteine (μmol/L) at screening, post-intervention (∼6-12 months later), 28 weeks gestation and at delivery in the mothers. B) Concentrations in the mother at delivery and in the cord blood of the baby are joined for the individual pairs. Concentrations of vitamin B12, holo-TC and folate were higher and those of total homocysteine lower in the cord blood compared to maternal concentrations at delivery. There was a highly significant association between maternal and cord blood concentrations of these four measurements (rho ∼ 0.7, p<0.001). * p<0.05, ** p<0.01, *** p<0.001 denotes significance of difference from the placebo; MMN, multiple micronutrients.

At delivery, compared to placebo, maternal vitamin B12 concentration was higher in the B12 alone group but not in the B12+MMN group. Holo-TC, folate and total homocysteine concentrations were similar in the three groups (Figure 2, Table 1). Vitamin B2 and B6 concentrations were highest in the mothers and in the cord blood of B12+MMN group. Maternal concentrations at delivery and cord blood concentrations of vitamin B12 (rho=0.716, p= <0.001), holo-TC (rho=0.668, p=<0.001), folate (rho=0.711, p=<0.001) and total homocysteine (rho=0.681, p=<0.001) were significantly correlated (Figure 2B). Cord blood hemoglobin concentration and red cell count were highest in the group supplemented with B12 alone. The total leucocyte count and individual counts of granulocytes, lymphocytes, monocytes, and platelets were similar in the 3 groups.

### RNA sequencing and differential gene expression analysis

Mononuclear cells from 88 cord blood samples were subjected to RNA sequencing. Each sample generated approximately 50 million reads. Out of the 88 samples, 9 samples showed poor alignment against the reference genome and were not considered for the downstream analysis. Our final analysis was therefore, on 79 samples. All samples showed comparable assignment to genes for generation of count matrix. The count matrix was used for differential expression analysis.

Samples showed clear separation based on the sex of the child on principal component analysis (PCA), however, no separation was observed based on any other covariate or the intervention (Figure 3A-3B). Differential expression analysis between the B12+MMN group and placebo resulted in identification of 75 differentially expressed genes (DEGs) (Figure 3C, Table 3, and Supplementary Table 3).

**Table 3:**
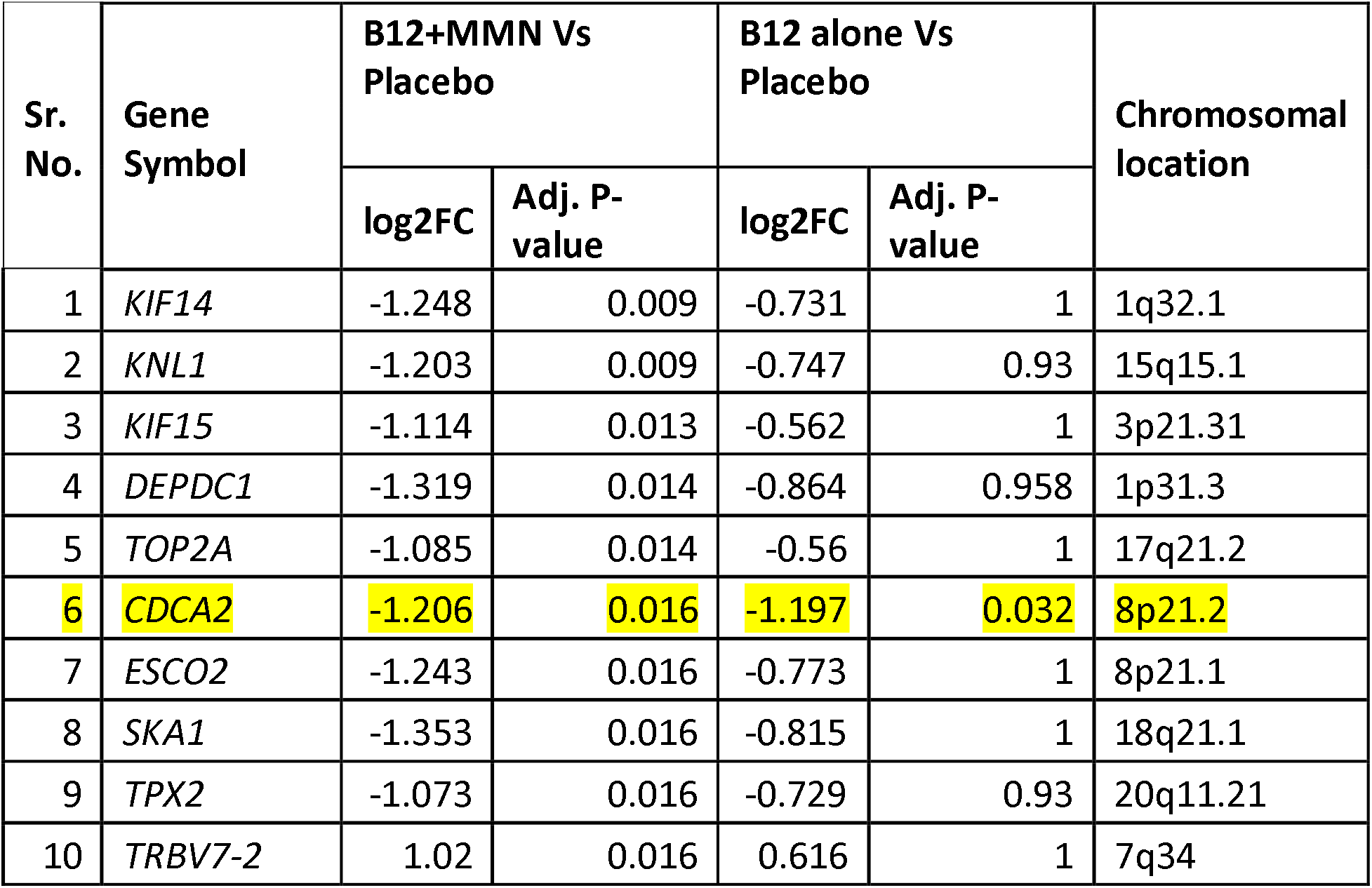
Top 10 genes that are differentially expressed between Placebo and B12+MMN groups.

**Figure 3:**
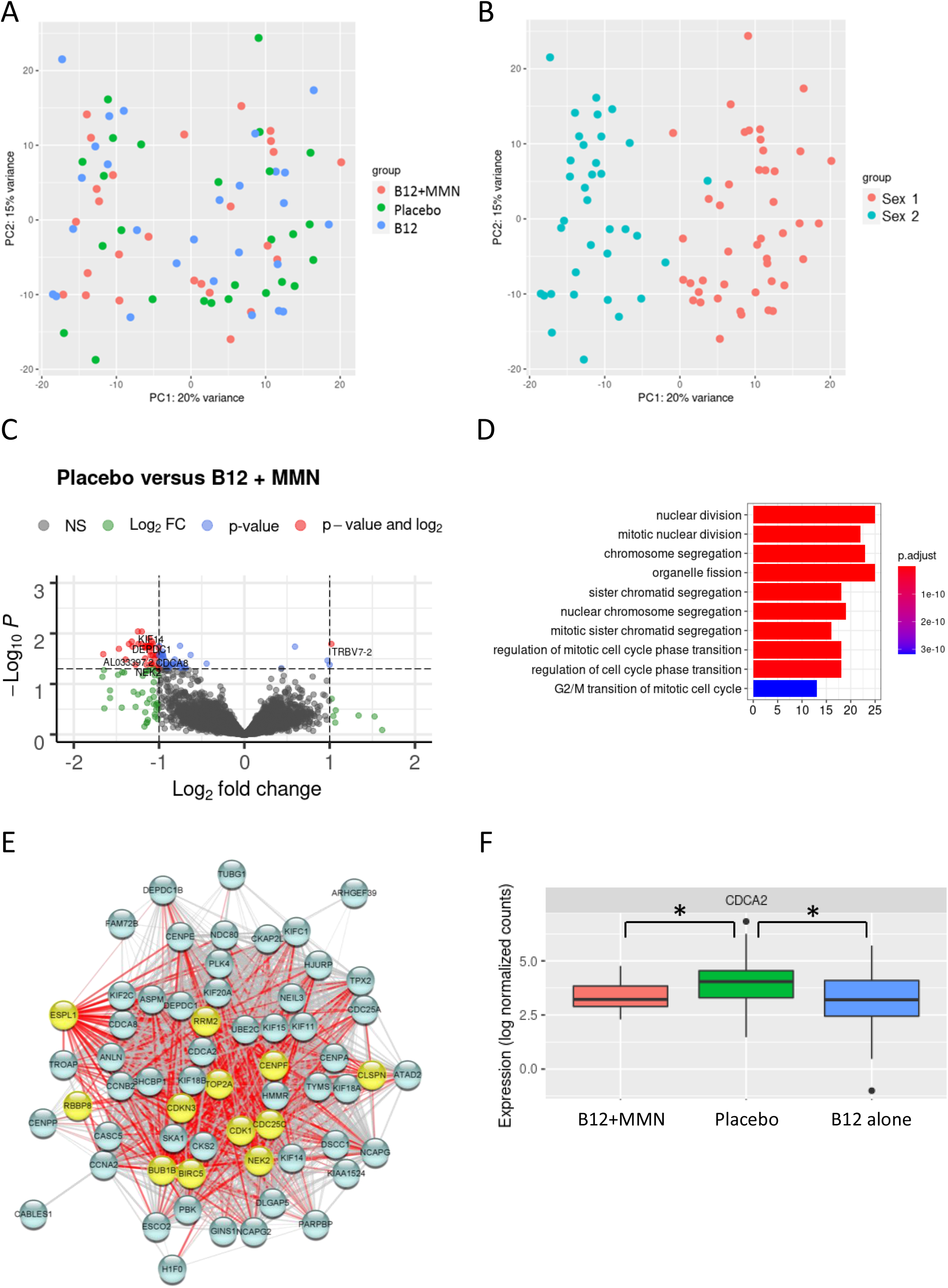
Gene enrichment analysis of genes differentially expressed between B12+MMN group and placebo. PCA plot highlighting A) intervention groups and B) sex of the child. C) Volcano plot for differential expression analysis. D) Gene enrichment and C) Network analysis of 75 differentially expressed genes highlighting negative regulators of cell cycle. D) Expression plot for CDCA2 in the three intervention groups (* adj P-value < 0.05); MMN, multiple micronutrients.

We used clusterProfiler and gProfiler to identify the top biological processes, pathways, and phenotypes affected by the DEGs (Figure 3D and Supplementary Figure 1). Participants supplemented with B12+MMN showed lower expression of genes associated with mitotic processes such as chromosome segregation, nuclear division, and G2/M phase transition as compared to the placebo group. To understand the gene-networking of the 75 DEGs, we performed analysis using STRING. Of the 75 DEGs, 71 formed a network with 12 showing association with the biological process ‘negative regulation of cell cycle’ (Figure 3E). Cell proliferation markers such as Cyclin D1 *(CCND1), PCNA*, and Ki-67 (*MKI67*) did not show significant differences in gene expression; however, pathway analysis identified enrichment of Cell Cycle Checkpoints and DNA damage response pathways. Enrichment of human phenotype terms Sloping forehead and Decreased head circumference was also observed. The 75 genes differentially expressed between B12+MMN and placebo also showed similar trend in expression in the B12 alone group but did not reach statistical significance (Supplementary Table 3). The B12 alone group showed two differentially expressed genes [Fructose Bis-phosphatase 2 (*FBP2*) and Cell Division Cycle Associated 2 (*CDCA2*)] compared to placebo of which, *CDCA2* was common to the B12+MMN group (Figure 3F). No significantly differentially expressed genes were observed between the two active intervention groups.

## Discussion

In this communication, we report the effect on the offspring’s cord blood transcriptome of a micronutrient intervention in mothers, started before conception (in adolescent age) and continued until the delivery of the child. The PRIYA trial is based in 6 villages near Pune and was guided by findings in the PMNS which showed: 1) a high prevalence of vitamin B12 deficiency in this predominantly vegetarian population^13, 14^ 2) that higher maternal total homocysteine concentration during pregnancy was associated with lower offspring birth weight^15^, 3) that low maternal vitamin B12 and normal to high folate status in pregnancy was associated with increased risk of diabesity in the child^14^, and 4) that lower maternal vitamin B12 and folate nutrition was associated with poorer neurocognitive performance in the child^27^. In the PRIYA trial, we supplemented adolescent participants from the PMNS with either vitamin B12 alone or with multi-micronutrients and milk protein, or only placebo. The vitamin B12+MMN intervention resulted in differential expression of 75 genes in the cord blood compared to the placebo group. Vitamin B12 alone influenced the expression of only a few genes, though the pattern of gene expression was similar in the two intervention groups. The differentially expressed genes in the B12+MMN group predominantly influenced mitosis related processes such as G2/M phase transition, chromosomal segregation, and nuclear division. B12 alone influenced transcription of *FBP2* and *CDCA2* genes of which *CDCA2* is common to B12+MMN group and involved in chromosomal segregation. To our knowledge, this is the first report of changes in the cord blood transcriptome after a maternal nutritional intervention initiated much prior to conception. Similar studies in the literature have mostly reported changes in DNA methylation in the cord blood but not gene expression. These include, response to maternal vitamin B12 supplementation^28-32^ or folic acid supplementation^33-38^. Effects of maternal micronutrient supplements other than vitamin B12 and folate on DNA methylation in the offspring have also been reviewed^39, 40^.

Nutritional deficiencies were common in the participants of the PMNS as reflected in low BMI of mothers, relatively low intake of calories, proteins and micronutrients from a predominantly vegetarian diet, high prevalence of anemia, low levels of vitamin B12 and ferritin, and low birth weight of the children^13, 14^. The villages were drought-prone, and the population had a low socio-economic status. Our intervention was prompted by our findings in this cohort which are summarized in the previous paragraph. In preparation for this intervention, we investigated and confirmed that absorption of a low dose oral vitamin B12 (2 mcg and 10 mcg) was adequate in this population, thus supporting the thesis that low vitamin B12 status was largely attributable to a low dietary intake^41^. We then performed a one-year pilot intervention to establish feasibility of a community-based trial and to decide on a dose of vitamin B12 which was physiological and effective in improving one carbon metabolism^42^. On this background, supplementation in the PRIYA trial achieved a demonstrable increase in circulating concentrations of vitamin B12, holo-TC, B2, and B6 in the B12 + MMN group and of vitamin B12 and holo-TC in the B12 alone group during the pre-conceptional period. The difference from the placebo group persisted in pregnancy even though there was the expected physiological fall in the levels of vitamin B12 and holo-TC. Improved vitamin status resulted in an improvement in maternal one-carbon metabolism, seen in a significant fall in plasma total homocysteine concentrations. In the B12+MMN group, though not measured, we expect an additional improvement in nutrition of vitamins A, D, and E, and other micronutrients (Zn etc.) given the contents of the UNIMMAP capsules (Supplementary table 1). High folate concentrations in pregnancy are attributable to iron and folic acid supplementation in all 3 groups as per Government of India approved dosage (60 mg elemental iron and 500 μg folic acid daily), thus it was also seen in the placebo group. It is interesting that the concentrations of the vitamins were higher in the cord blood compared to the maternal blood, suggesting active placental transfer. Overall, our intervention translated into better micronutrient nutrition of the ovum before conception and of the conceptus throughout the pregnancy. A relative lack of difference in micronutrient concentrations between intervention and placebo groups in the cord blood may be contributed by altered placental transport, altered binding to carrier proteins, and differential utilisation by the baby^43, 44^.

An inspection of the one-carbon metabolic cycle (Figure 4) and its regulation by B-complex vitamins suggests that mothers and babies who received vitamin B12+MMN would have a more comprehensive effect on nucleotide synthesis, and DNA and RNA metabolism compared to those who received vitamin B12 alone. Higher hemoglobin and red blood cell count in the vitamin B12 alone group may be related to higher levels of vitamin B12 and holo-TC in this group and their specific effect on haemopoietic tissue.

**Figure 4:**
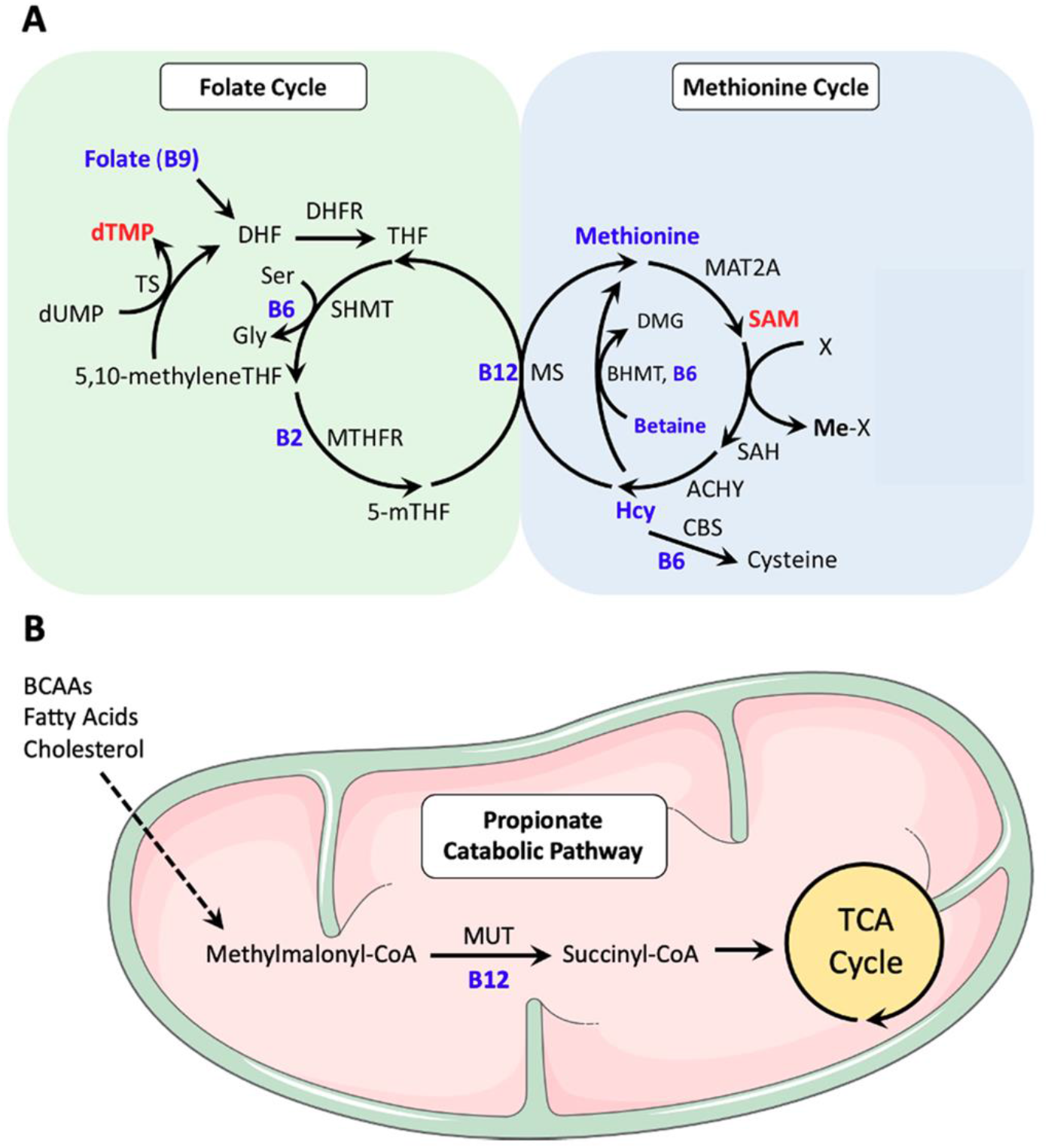
One-Carbon Metabolism and PRIYA trial. A) One carbon metabolism is a central metabolic pathway combining folate cycle and methionine cycle. Folate cycle generates nucleotides which help DNA synthesis and repair while methionine cycle generates methionine and S-adenosyl methionine (SAM). Methionine is a component of all cellular proteins and SAM methylates a wide range of cellular molecules including DNA (an epigenetic mechanism), lipids (important in nervous system), and proteins. Folate cycle is influenced by a number of B complex vitamins (B2, B6, B9 and B12) as is the methionine cycle (B6, B12). Dietary proteins provide the amino acid serine which is the primary donor of methyl groups, and other amino acids which get incorporated in protein molecules. B) Vitamin B12 has an additional influence on mitochondrial energy metabolism through its action as a cofactor to mutase enzyme which converts methylmalonic acid to succinic acid (Adapted from Lyon et al., 2020^53^).

A detailed analysis of the transcriptomic differences suggested that though the overall pattern of differential expression was similar in the two intervention groups, the effect size was greater in the B12+MMN group. Thus, 75 genes were differentially expressed in the B12+MMN group but only two genes (*FBP2* and *CDCA2*) in the B12 alone group. Seventy-one of these 75 differentially expressed genes were closely networked and twelve of them showed a negative association with regulation of cell cycle. The affected biological processes included cell cycle G2/M phase transition, chromosome segregation, nuclear division and related. However, a targeted expression analysis of cell proliferation markers such as cyclin D1, Minichromosome Maintenance Complex Component 2 (*MCM2*), Proliferating Cell Nuclear Antigen (*PCNA*) and Ki-67^44^, didn’t show any significant difference in any of the intervention groups compared to the placebo group. Intriguingly, cell cycle checkpoint and DNA repair related genes showed higher expression in the placebo group.

We propose that low concentrations of micronutrients in the placebo group resulted in hypomethylation of the relevant genes and consequently increased the expression of genes involved in nucleic acid synthesis and cell division. However, micronutrient deficiency compromised the supply of nucleotides for DNA synthesis and replication resulting in activation of cell cycle checkpoint. Thus, genes related to DNA synthesis and cell division appear relatively under-expressed in the B12+MMN group because the intervention led to better micronutrient status influencing methylation status of those relevant genes. Interestingly, in another study of vitamin B12 and/or folic acid supplementation in 9 year old children, we reported differential DNA methylation of genes associated with G2/M checkpoint regulation^45^. The MMN supplementation also included vitamins A and D, and zinc which are known to affect gene expression in specific tissues^46-48^ and may have contributed to some of differentially expressed genes in the B12+MMN group. A review has summarized the recent exciting understanding of the role of core cellular metabolites (ATP, S-adenosyl methionine, acetyl-CoA, NAD/NADP and α-ketoglutarate) in the nuclear compartment to influence the epigenome; all these metabolites are likely to be influenced in our intervention^49^. Association of transcriptomic changes with aspects of facial and head development is intriguing and it will be of interest to see if they reflect in any phenotypic features during follow up of the children, especially in the neurocognitive domain.

Features of vitamin B12 deficiency are usually described in relation to pernicious anaemia (a genetic-immunologic condition causing absence of ‘intrinsic factor’ which is essential for vitamin B12 absorption). The biochemical and cellular changes include: block in deoxyuridine to thymidine conversion leading to disturbed DNA synthesis and repair, prolongation of S-phase, maturation arrest (G2/M transition) and failure of cell division^50^. There is a remarkable similarity in these disturbances and our findings. We could not find any reports of transcriptomic alterations in this condition. Two studies investigated the effect of cobalamin deficiency on nerve cells in culture: there was slower proliferation and accelerated differentiation of neuroblastoma cells^51^ and impaired DNA synthesis, prolonged G2/M phase transition leading to cell growth without cell division in human astrocytes^52^. Again, no transcriptomic data was available.

## Strengths and Limitations

A major strength of our intervention is that it was guided by past nutritional observations in the same cohort and that it was truly pre-conceptional (from adolescent age). Thus, our trial had the potential to influence peri-conceptional epigenetic reprogramming. Use of ‘physiological’ doses of micronutrients was based on the successful results of an absorption study and a pilot trial^42^ which made it public health relevant. The limitations include that these preliminary results are on a relatively small number. The limited power could also have contributed to a lack of difference in the cord blood measurements of some of the micronutrients and in body size measurements of the babies in the three groups. Another limitation is that the transcriptomic measurements are limited only to cord blood mononuclear cells which may not reflect changes in other tissues and those earlier in the pregnancy. Such invasive studies are difficult in humans for ethical reasons. It should be noted that the transcriptome analysis using cord blood mononuclear cells acts as a surrogate to gain insights into the multitude of changes in the gene expression profiles resulted due to the in-utero programming following the nutritional intervention.

In conclusion, we provide the first evidence of the influence of human maternal pre-conceptional micronutrient intervention on the gene expression profile of the cord blood at delivery. Supplementation with vitamin B12 and other micronutrients influenced vital cellular processes such as cell cycle regulation, mitotic division, and growth of facial and head structures. These gene expression changes provide plausible mechanisms mediating intrauterine programming of future health and disease. In subsequent papers we intend to report the results of cord blood methylome, proteome, and metabolome and use a systems approach to allow a better understanding of the full range of ‘OMIC’ changes in response to micronutrient intervention. Follow up studies in children will provide an opportunity to investigate the association of these alterations with outcomes of body size and composition, metabolism, and neurocognitive function.

## Data availability

The datasets generated for this study can be found in NCBI SRA: PRJNA756634. Funding

The work was supported by research grant titled “Centre of Excellence in Fetal Programming” from Department of Biotechnology (DBT), Government of India (CEIB-BT/PR12629/MED/97/364/2016) and Indian Council of Medical Research (ICMR) – Medical Research Council (MRC), UK grant (No.58/1/8/ICMR-MRC/2009-NCD-II.

## Data Availability

The datasets generated for this study can be found in NCBI SRA: PRJNA756634.

https://www.ncbi.nlm.nih.gov/bioproject/PRJNA756634/

## Acknowledgements

Ayush Madhok, Indumathi Patta, and Vipul V. Wagh thank CSIR-UGC, Government of India for research fellowships. Satyajeet P. Khare is a beneficiary of the DST SERB SRG grant (SRG/2020/001414). The authors thank Drs Ameya Sathe, Ankitha Shetty for their help in processing cord blood samples, Saurabh Pradhan for help with RNA-seq, Neelam Memane and Rajashree Kamat for laboratory assays, Aboli Bhalerao and Madhura Deshmukh, field staff for their help in the PRIYA study, Mrs. Pallavi Yajnik and Rasika Ladkat for administrative help.

## Author contributions

Chittaranjan S. Yajnik and Caroline Fall designed the PRIYA trial. Kalyanaraman Kumaran contributed to running the trial. Indumathi Patta assisted in optimization of cell isolation and RNA preparation. Satyajeet P. Khare and Ayush Madhok performed RNA sequencing and analysis, Satyajeet P. Khare wrote the manuscript. Krishna K. Sukla, Vipul Wagh, and Pooja S. Kunte helped in data analysis and drafting the manuscript. Giriraj R. Chandak and Utpal Tatu contributed to the discussion and interpretations. Deepa Raut and Dattatray Bhat assisted in sample collection. Sanjeev Galande designed the experiments. Chittaranjan S. Yajnik and Sanjeev Galande discussed and interpreted the results, supported the research, and wrote the manuscript. All authors read and approved the manuscript.

