## Supplementary Table 1 for "Differential expression of genes influencing mitotic processes in cord blood mononuclear cells after a pre-conceptional micronutrient-based randomized controlled trial: Pune Rural Intervention in Young Adolescents (PRIYA)"

**Supplementary table 1**: Contents of the multiple micronutrient (MMN) capsule in the PRIYA trial and comparison with United Nations International Multiple Micronutrient Antenatal Preparation (UNIMMAP).

| **Contents** | **UNIMMAP (1999)** | **PRIYA** | |
| --- | --- | --- | --- |
|  |  | **B12 + MMN** | **B12 alone** |
| **Vitamin A (μg)** | 800 | 300 | - |
| **Vitamin D (IU)** | 200 | 100 | - |
| **Vitamin E (mg)** | 10 | 5 | - |
| **Vitamin C (mg)** | 70 | 20 | - |
| **Vitamin B1 (mg)** | 1.4 | 0.75 | - |
| **Vitamin B2 (mg)** | 1.4 | 0.9 | - |
| **Vitamin B3 (mg)** | 18 | 10 | - |
| **Vitamin B6 (mg)** | 1.9 | 0.5 | - |
| **Vitamin B12 (μg)** | 2.4 | 1 | 1 |
| **Zinc (mg)** | 15 | 6 | - |
| **Copper (mg)** | 2 | 1 | - |
| **Selenium (μg)** | 65 | 20 | - |
| **Iodine (μg)** | 150 | 75 | - |

**Note**: Dose in PRIYA trial was 2 capsules/day. Placebo group capsule did not contain micronutrients. Iron and folic acid tablet was given separately to all groups as per the Government of India guidelines.
