## Supplementary Table 2 for "Differential expression of genes influencing mitotic processes in cord blood mononuclear cells after a pre-conceptional micronutrient-based randomized controlled trial: Pune Rural Intervention in Young Adolescents (PRIYA)"

**Supplementary Table 2**: Following tests were performed at different time points in the trial

| Analyte | Timepoint | Test | Instrument | Comment |
| --- | --- | --- | --- | --- |
| Complete Blood Count | Baseline, 18y, 28 weeks gestation, delivery, cord blood | Whole blood collected and analysed on the day of visit | Beckman Coulter analyser (AC.T diffTM Analyzer, Miami, Florida, USA). | - |
| Vitamin B12 | Baseline, 18 year and 28 weeks gestation | Microbiological assay on plasma using a colistin sulfate-resistant strain of L. Leichmanii | - | This is our standard research assay. |
|  |  |  |  | Sensitivity: 50 pmol/L |
|  |  |  |  | CV: <8% |
|  |  |  |  | Antibiotic treatment interferes with this assay. |
|  | Delivery and cord blood | Electro Chemiluminescence Immuno Assay (ECLIA) on plasma | Cobas e-411 analyzer, ROCHE diagnostics GmbH, Sandhofer Strasse 116, Mannheim, Germany | We used this assay because women undergoing caesarean section received antibiotics before delivery which interferes with microbial assay. |
|  |  |  |  | Correlation coefficient with Microbiological assay r = 0.958 by paired t-test |
|  |  |  |  | Sensitivity: 62 pmol/L |
|  |  |  |  | CV: 7% |
| Holo-TC (Holo-transcobalamin) | Baseline, 18y, 28 weeks gestation, delivery, cord blood | Chemiluminescent microparticle immunoassay (CMIA) on plasma, using anti-holotranscobalamin coated paramagnetic microparticles | ARCHITECT (ABBOTT-GmbH &amp; Co. KG Max-Planck-Ring2 65205 Wiesbaden Germany +49-6122-580) | Sensitivity: <= 5.0 pmol/L |
|  |  |  |  | CV: <= 8.5 % |
| Folate | Baseline, 18y, 28 weeks gestation, delivery, cord blood | Microbiological assay on plasma using a chloramphenicol-resistant strain of L. Casei | - | Sensitivity: 3nmol/L |
|  |  |  |  | CV: <8% |
| Total homocysteine | Baseline, 18y, 28 weeks gestation, delivery, cord blood | HPLC on plasma | PerkinElmer 200 Series, PerkinElmer, Shelton, CT, USA, using fluorescence detector | Sensitivity: 3 µmol/L |
|  |  |  |  | CV: <4% |
| Vitamin B2 | 18y, 28 weeks gestation, delivery, cord blood | Kits + HPLC on whole blood | Recipe Chemicals + Instruments GmbH, Munchen, Germany); HPLC (PerkinElmer 200 Series, PerkinElmer, Shelton, CT, USA) with fluorescence detector | Sensitivity: 10 µg/L |
|  |  |  |  | CV: 4% |
| Vitamin B6 | Baseline, 18y, 28 weeks gestation, delivery, cord blood | Kits + HPLC on plasma | Recipe Chemicals + Instruments GmbH, Munchen, Germany); HPLC (PerkinElmer 200 Series, PerkinElmer, Shelton, CT, USA) with fluorescence detector | Sensitivity: 0.4 µg/L |
|  |  |  |  | CV: 5.1% |
| Neonatal birth weight | At birth | Salter spring balance | - | - |
| Neonatal birth length | At birth | Portable pedobaby babymeter | ETS JMB, Brussels, Belgium | - |
| Neonatal skinfolds | At birth | Harpenden skinfold callipers | CMS Instruments, London, UK | - |
| Maternal weight | 28 weeks of gestation | Electronic weighing scales | ATCO Healthcare Ltd, Mumbai, India | - |
| Maternal height | 28 weeks of gestation | Wall-mounted Microtoise | CMS Instruments Ltd, London, UK | - |

NOTE:

1. Details of biochemical measurements are described in Yajnik CS, Deshpande SS, Jackson AA, et al. Vitamin B12 and folate concentrations during pregnancy and insulin resistance in the offspring: the Pune Maternal Nutrition Study. Diabetologia. 2008;51(1):29-38. doi:10.1007/s00125-007-0793-y

2. Details of anthropometric measuremetns are described in Yajnik CS, Fall CH, Coyaji KJ, et al. Neonatal anthropometry: the thin-fat Indian baby. The Pune Maternal Nutrition Study. Int J Obes Relat Metab Disord. 2003;27(2):173-180. doi:10.1038/sj.ijo.802219
