## Supplementary Table 3 for "Differential expression of genes influencing mitotic processes in cord blood mononuclear cells after a pre-conceptional micronutrient-based randomized controlled trial: Pune Rural Intervention in Young Adolescents (PRIYA)"

**Supplementary table 3**: List of genes that are differentially expressed between Placebo and B12+MMN groups.

| **Sr. No.** | **Gene Symbol** | **Placebo vs B12+MMN** | | **Placebo vs B12 alone** | | **Chromosomal location** |
| --- | --- | --- | --- | --- | --- | --- |
|  |  | **(Group2 Vs Group1)** | | **(Group2 Vs Group3)** | |  |
|  |  | **log2FC** | **Adj. P-value** | **log2FC** | **Adj. P-value** |  |
| 1 | *KIF14* | -1.248 | 0.009 | -0.731 | 1 | 1q32.1 |
| 2 | *KNL1* | -1.203 | 0.009 | -0.747 | 0.93 | 15q15.1 |
| 3 | *KIF15* | -1.114 | 0.013 | -0.562 | 1 | 3p21.31 |
| 4 | *DEPDC1* | -1.319 | 0.014 | -0.864 | 0.958 | 1p31.3 |
| 5 | *TOP2A* | -1.085 | 0.014 | -0.56 | 1 | 17q21.2 |
| 6 | *CDCA2* | -1.206 | 0.016 | -1.197 | 0.032 | 8p21.2 |
| 7 | *ESCO2* | -1.243 | 0.016 | -0.773 | 1 | 8p21.1 |
| 8 | *SKA1* | -1.353 | 0.016 | -0.815 | 1 | 18q21.1 |
| 9 | *TPX2* | -1.073 | 0.016 | -0.729 | 0.93 | 20q11.21 |
| 10 | *TRBV7-2* | 1.02 | 0.016 | 0.616 | 1 | 7q34 |
| 11 | *ASPM* | -1.091 | 0.018 | -0.751 | 1 | 1q31.3 |
| 12 | *CABLES1* | -1.137 | 0.018 | -0.273 | 1 | 18q11.2 |
| 13 | *CDKN3* | -1.11 | 0.018 | -0.44 | 1 | 14q22.2 |
| 14 | *CLSPN* | -0.998 | 0.018 | -0.544 | 1 | 1p34.3 |
| 15 | *NCAPG* | -1.164 | 0.018 | -0.665 | 1 | 4p15.31 |
| 16 | *NCAPG2* | -0.755 | 0.018 | -0.402 | 1 | 7q36.3 |
| 17 | *NEIL3* | -1.118 | 0.018 | -0.416 | 1 | 4q34.3 |
| 18 | *BICDL1* | 0.594 | 0.018 | 0.188 | 1 | 12q24.23 |
| 19 | *CDK1* | -1.311 | 0.018 | -0.631 | 1 | 10q21.2 |
| 20 | *HMMR* | -1.116 | 0.018 | -0.583 | 1 | 5q34 |
| 21 | *PRSS21* | -1.155 | 0.018 | -0.719 | 1 | 16p13.3 |
| 22 | *ESPL1* | -1.164 | 0.019 | -0.946 | 0.698 | 12q13.13 |
| 23 | *KIF20A* | -1.091 | 0.019 | -0.715 | 1 | 5q31.2 |
| 24 | *SHCBP1* | -1.006 | 0.019 | -0.594 | 1 | 16q11.2 |
| 25 | *PBK* | -1.47 | 0.02 | -0.662 | 1 | 8p21.1 |
| 26 | *CENPF* | -0.96 | 0.023 | -0.548 | 1 | 1q41 |
| 27 | *DEPDC1B* | -1.106 | 0.023 | -0.735 | 1 | 5q12.1 |
| 28 | *AL033397.2* | -1.654 | 0.026 | -1.103 | 1 | NA |
| 29 | *KIF11* | -0.977 | 0.026 | -0.543 | 1 | 10q23.33 |
| 30 | *UBE2C* | -0.959 | 0.026 | -0.526 | 1 | 20q13.12 |
| 31 | *ATAD2* | -0.63 | 0.027 | -0.281 | 1 | 8q24.13 |
| 32 | *CDCA8* | -1.033 | 0.027 | -0.815 | 0.811 | 1p34.3 |
| 33 | *CKAP2L* | -1.028 | 0.027 | -0.718 | 1 | 2q14.1 |
| 34 | *H1F0* | -0.953 | 0.027 | -0.55 | 1 | 22q13.1 |
| 35 | *ANLN* | -0.996 | 0.027 | -0.445 | 1 | 7p14.2 |
| 36 | *FAM72B* | -0.998 | 0.027 | -0.799 | 0.811 | 1p11.2 |
| 37 | *KIFC1* | -1.075 | 0.027 | -0.625 | 1 | 6p21.32 |
| 38 | *TROAP* | -1.098 | 0.027 | -0.925 | 0.722 | 12q13.12 |
| 39 | *CCNA2* | -0.949 | 0.029 | -0.574 | 1 | 4q27 |
| 40 | *CENPA* | -1.163 | 0.031 | -0.6 | 1 | 2p23.3 |
| 41 | *CENPE* | -0.966 | 0.031 | -0.643 | 1 | 4q24 |
| 42 | *RRM2* | -1.048 | 0.031 | -0.603 | 1 | 2p25.1 |
| 43 | *KIF18B* | -1.393 | 0.033 | -0.633 | 1 | 17q21.31 |
| 44 | *PARPBP* | -0.939 | 0.033 | -0.363 | 1 | 12q23.2 |
| 45 | *TUBG1* | -0.816 | 0.033 | -0.808 | 0.181 | 17q21.31 |
| 46 | *TRBV6-2* | 0.974 | 0.034 | 0.628 | 1 | 7q34 |
| 47 | *BUB1B* | -0.903 | 0.039 | -0.401 | 1 | 15q15.1 |
| 48 | *HJURP* | -1 | 0.039 | -0.591 | 1 | 2q37.1 |
| 49 | *CIP2A* | -0.732 | 0.04 | -0.377 | 1 | 3q13.13 |
| 50 | *RBBP8* | -0.442 | 0.04 | -0.2 | 1 | 18q11.2 |
| 51 | *BIRC5* | -1.089 | 0.04 | -0.554 | 1 | 17q25.3 |
| 52 | *CENPP* | -0.802 | 0.04 | -0.43 | 1 | 9q22.31 |
| 53 | *AC091057.1* | -0.845 | 0.041 | -0.323 | 1 | NA |
| 54 | *DLGAP5* | -1.089 | 0.041 | -0.625 | 1 | 14q22.3 |
| 55 | *DSCC1* | -0.989 | 0.041 | -0.817 | 0.811 | 8q24.12 |
| 56 | *PLK4* | -0.753 | 0.041 | -0.432 | 1 | 4q28.1 |
| 57 | *AC093890.2* | -0.851 | 0.042 | -0.481 | 1 |  |
| 58 | *CDC25A* | -1.057 | 0.042 | -0.808 | 1 | 3p21.31 |
| 59 | *AC027237.1* | -0.864 | 0.042 | -0.463 | 1 | NA |
| 60 | *ADD2* | -0.922 | 0.042 | -0.678 | 1 | 2p13.3 |
| 61 | *ARHGEF39* | -0.797 | 0.042 | -0.492 | 1 | 9p13.3 |
| 62 | *CD34* | -0.916 | 0.042 | -0.595 | 1 | 1q32.2 |
| 63 | *CDC25C* | -1.146 | 0.042 | -0.501 | 1 | 5q31.2 |
| 64 | *KIF2C* | -0.941 | 0.042 | -0.61 | 1 | 1p34.1 |
| 65 | *MIR4697HG* | 0.999 | 0.042 | 0.691 | 1 | 11q25 |
| 66 | *NEK2* | -1.275 | 0.042 | -0.861 | 1 | 1q32.3 |
| 67 | *CCNB2* | -1.019 | 0.047 | -0.583 | 1 | 15q22.2 |
| 68 | *CKS2* | -0.729 | 0.048 | -0.29 | 1 | 9q22.2 |
| 69 | *NDC80* | -0.75 | 0.048 | -0.57 | 1 | 18p11.32 |
| 70 | *WDR34* | -0.688 | 0.048 | -0.416 | 1 | 9q34.11 |
| 71 | *KIF18A* | -0.886 | 0.048 | -0.605 | 1 | 11p14.1 |
| 72 | *TYMS* | -0.91 | 0.048 | -0.503 | 1 | 18p11.32 |
| 73 | *CRELD1* | 0.434 | 0.049 | 0.129 | 1 | 3p25.3 |
| 74 | *GCLM* | -0.702 | 0.049 | -0.435 | 1 | 1p22.1 |
| 75 | *GINS1* | -1.001 | 0.05 | -0.417 | 1 | 20p11.21 |
