## Supplementary Figure 1 for "Differential expression of genes influencing mitotic processes in cord blood mononuclear cells after a pre-conceptional micronutrient-based randomized controlled trial: Pune Rural Intervention in Young Adolescents (PRIYA)"

REAC

| Term name | Term ID | $p_{adj}$ |
|---|---|---|
| Cell Cycle | REAC:R-HSA-16... | $3.134\times10^{-31}$ |
| Cell Cycle, Mitotic | REAC:R-HSA-69... | $3.760\times10^{-28}$ |
| Mitotic Prometaphase | REAC:R-HSA-68... | $1.828\times10^{-16}$ |
| Cell Cycle Checkpoints | REAC:R-HSA-69... | $2.949\times10^{-16}$ |
| M Phase | REAC:R-HSA-68... | $6.334\times10^{-16}$ |
| Resolution of Sister Chromatid Cohesion | REAC:R-HSA-25... | $1.047\times10^{-13}$ |
| Mitotic Spindle Checkpoint | REAC:R-HSA-69... | $7.437\times10^{-13}$ |
| Mitotic Anaphase | REAC:R-HSA-68... | $1.374\times10^{-12}$ |
| Mitotic Metaphase and Anaphase | REAC:R-HSA-25... | $1.473\times10^{-12}$ |
| Amplification of signal from unattached kinetochores via a ... | REAC:R-HSA-14... | $3.336\times10^{-12}$ |
| Amplification of signal from the kinetochores | REAC:R-HSA-14... | $3.336\times10^{-12}$ |
| Separation of Sister Chromatids | REAC:R-HSA-24... | $4.229\times10^{-11}$ |
| EML4 and NUDC in mitotic spindle formation | REAC:R-HSA-96... | $5.246\times10^{-11}$ |
| RHO GTPases Activate Formins | REAC:R-HSA-56... | $4.019\times10^{-10}$ |
| RHO GTPase Effectors | REAC:R-HSA-19... | $5.553\times10^{-8}$ |
| Kinesins | REAC:R-HSA-98... | $1.154\times10^{-7}$ |
| G2/M Transition | REAC:R-HSA-69... | $5.007\times10^{-7}$ |
| Mitotic G2-G2/M phases | REAC:R-HSA-45... | $5.580\times10^{-7}$ |
| Signaling by Rho GTPases | REAC:R-HSA-19... | $3.579\times10^{-6}$ |
| Signaling by Rho GTPases, Miro GTPases and RHOBTB3 | REAC:R-HSA-97... | $4.922\times10^{-6}$ |
| COPI-dependent Golgi-to-ER retrograde traffic | REAC:R-HSA-68... | $6.715\times10^{-6}$ |
| Factors involved in megakaryocyte development and platel... | REAC:R-HSA-98... | $3.076\times10^{-5}$ |
| Cyclin A/B1/B2 associated events during G2/M transition | REAC:R-HSA-69... | $3.666\times10^{-5}$ |
| Golgi-to-ER retrograde transport | REAC:R-HSA-88... | $6.805\times10^{-5}$ |
| Polo-like kinase mediated events | REAC:R-HSA-15... | $2.227\times10^{-4}$ |
| AURKA Activation by TPX2 | REAC:R-HSA-88... | $3.593\times10^{-4}$ |
| Intra-Golgi and retrograde Golgi-to-ER traffic | REAC:R-HSA-68... | $1.621\times10^{-3}$ |
| Mitotic G1 phase and G1/S transition | REAC:R-HSA-45... | $2.042\times10^{-3}$ |
| G0 and Early G1 | REAC:R-HSA-15... | $2.347\times10^{-3}$ |
| Condensation of Prometaphase Chromosomes | REAC:R-HSA-25... | $6.105\times10^{-3}$ |
| MHC class II antigen presentation | REAC:R-HSA-21... | $8.845\times10^{-3}$ |
| APC:Cdc20 mediated degradation of cell cycle proteins prio... | REAC:R-HSA-17... | $9.332\times10^{-3}$ |
| APC/C:Cdc20 mediated degradation of mitotic proteins | REAC:R-HSA-17... | $1.063\times10^{-2}$ |
| Activation of APC/C and APC/C:Cdc20 mediated degradatio... | REAC:R-HSA-17... | $1.132\times10^{-2}$ |
| G1/S Transition | REAC:R-HSA-69... | $1.210\times10^{-2}$ |
| Chromosome Maintenance | REAC:R-HSA-73... | $1.765\times10^{-2}$ |
| Regulation of PLK1 Activity at G2/M Transition | REAC:R-HSA-25... | $1.827\times10^{-2}$ |
| Regulation of mitotic cell cycle | REAC:R-HSA-45... | $2.159\times10^{-2}$ |
| APC/C-mediated degradation of cell cycle proteins | REAC:R-HSA-17... | $2.159\times10^{-2}$ |
| Phosphorylation of proteins involved in the G2/M transition... | REAC:R-HSA-17... | $2.495\times10^{-2}$ |
| S Phase | REAC:R-HSA-69... | $3.997\times10^{-2}$ |

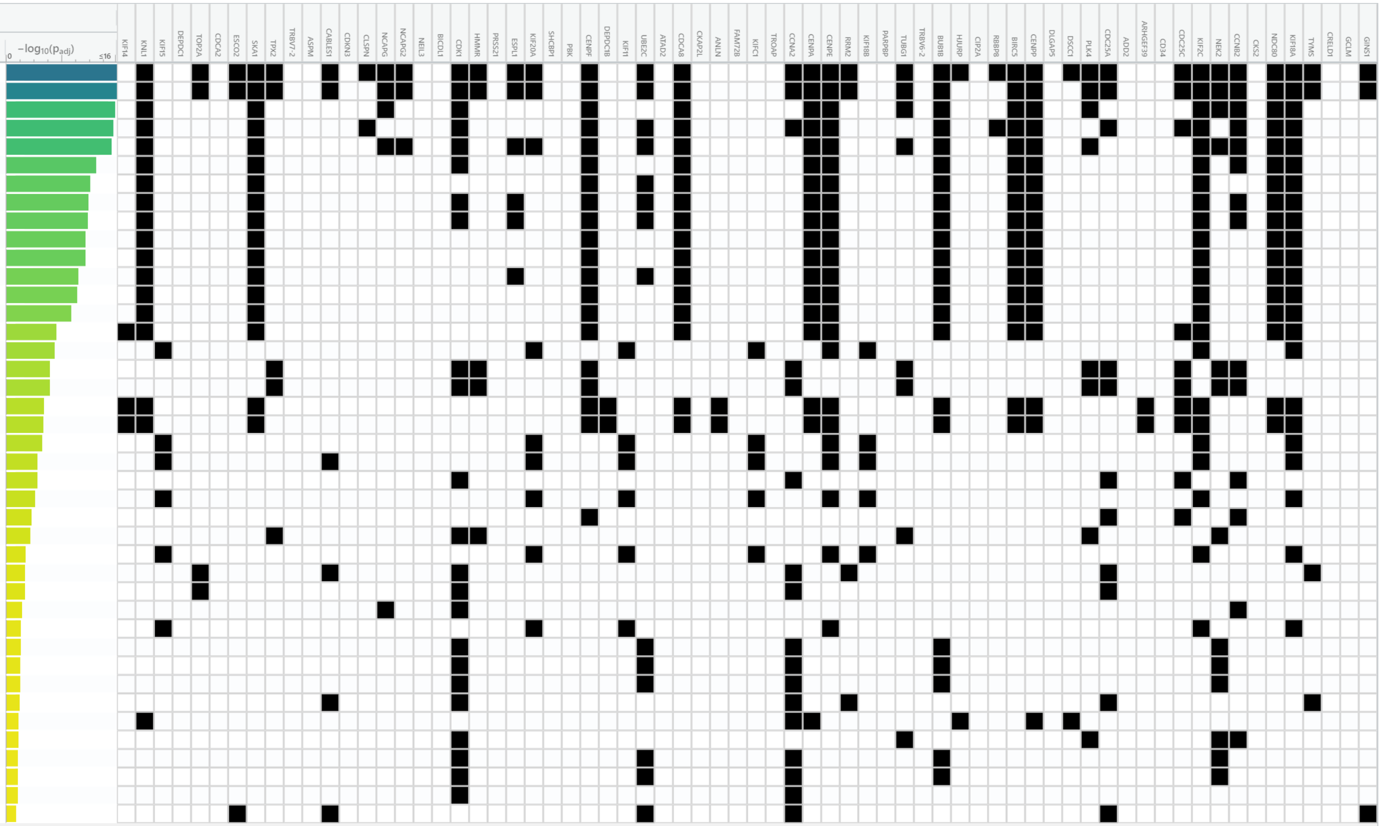


1 to 41 of 41 Page 1 of 1

WP

| Term name | Term ID | $p_{adj}$ |
|---|---|---|
| Retinoblastoma gene in cancer | WP:WP2446 | $7.032\times10^{-9}$ |
| Gastric cancer network 1 | WP:WP2361 | $1.276\times10^{-5}$ |
| Gastric cancer network 2 | WP:WP2363 | $1.166\times10^{-3}$ |
| Cell cycle | WP:WP179 | $1.186\times10^{-3}$ |
| Regulation of sister chromatid separation at the metaphase... | WP:WP4240 | $4.359\times10^{-3}$ |
| DNA damage response | WP:WP707 | $2.214\times10^{-2}$ |
| DNA IR-damage and cellular response via ATR | WP:WP4016 | $4.111\times10^{-2}$ |
| Cohesin complex - Cornelia de Lange syndrome | WP:WP5117 | $4.823\times10^{-2}$ |

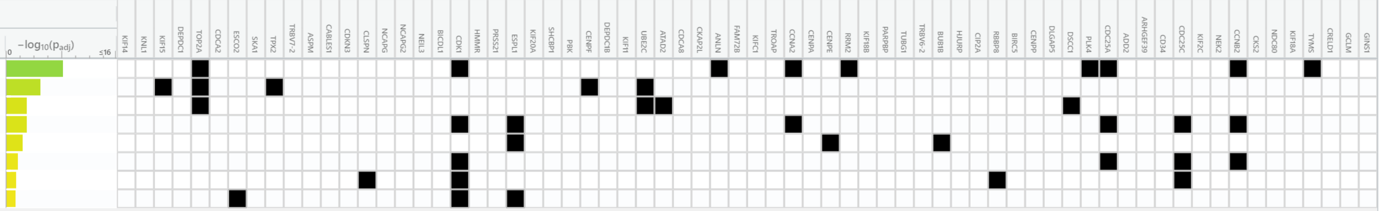

1 to 8 of 8 Page 1 of 1

HP

| Term name | Term ID | $p_{adj}$ |
|---|---|---|
| Absent earlobe | HP:0000387 | $4.660\times10^{-4}$ |
| Sloping forehead | HP:0000340 | $6.104\times10^{-3}$ |
| Simplified gyral pattern | HP:0009879 | $8.935\times10^{-3}$ |
| Microcephaly | HP:0000252 | $2.271\times10^{-2}$ |
| Abnormality of globe size | HP:0100887 | $2.286\times10^{-2}$ |
| Decreased head circumference | HP:0040195 | $2.634\times10^{-2}$ |
| Aplasia/Hypoplasia of the earlobes | HP:0009906 | $4.257\times10^{-2}$ |
| Slanting of the palpebral fissure | HP:0200006 | $4.557\times10^{-2}$ |

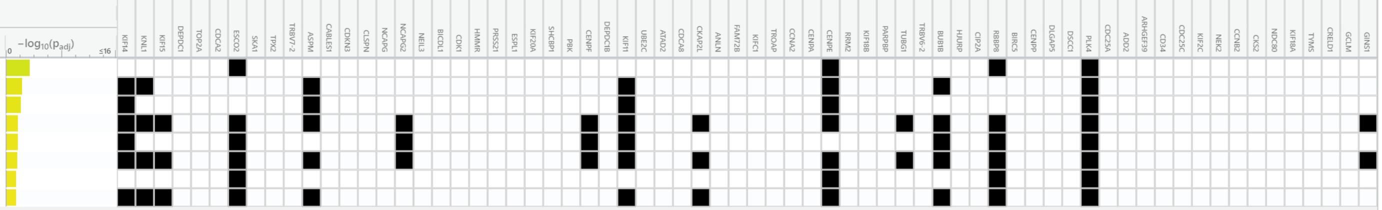

1 to 8 of 8 Page 1 of 1
